# Myoconjunctival Enucleation Study (MES): Outcome of Myoconjunctival enucleation technique with polymethyl Metha acrylate (PMMA) implant with custom made prosthesis (CMP) in tertiary eye care center in Nepal

**DOI:** 10.1101/2025.03.13.25323811

**Authors:** Hom Bahadur Gurung, Purnima Rajkarnikar Sthapit, Malita Amatya, Dikshya Bista, Sushant Adiga, Manish Poudel, Raba Thapa, Rohit Saiju

## Abstract

**Purpose:** This study aims to evaluate the average motility of the implant and custom-made prosthesis after MES, the surgical duration, and the complications encountered during the procedure at a tertiary eye hospital.

**Methods:** This was a prospective, non-randomized interventional study. The muscles were sutured to the fornices before the enucleation of the eyeball to minimize surgical time and the need for additional sutures. 35 consecutive patients meeting the inclusion criteria were enrolled in the study. Data on surgery time, complication rates, and implant/prosthesis motility were recorded and analyzed.

**Results:** The mean surgery duration was 39.3 ± 3.76 minutes. The mean movement of the PMMA implant was 2.34 mm in elevation, 2.25 mm in depression, 2.74 mm in abduction, and 2.71 mm in adduction. The mean movement of the custom-made prosthesis was 2.45 mm in elevation, 2.62 mm in depression, 2.74 mm in adduction, and 2.82 mm in abduction. Complications were minimal: one intraoperative superior rectus slippage, few cases with early postoperative nausea, vomiting, and headache, all resolving within a week. At six weeks, one case each of suture granuloma and shallow inferior fornix was noted and managed successfully. Patient satisfaction was high (91.94%), with a mean score of 4.25 ± 0.65. Minimal dissatisfaction was linked to low motility or mild socket contraction.

**Conclusion:** Myoconjunctival enucleation offers favorable implant and prosthesis motility, a relatively short surgical duration, and minimal complications. High patient satisfaction further supports its efficacy as a reliable enucleation technique.

## Introduction

Enucleation procedures are on the rise in many tertiary centres, primarily due to increased referrals for conditions such as intraocular tumors, phthisis bulbi, end-stage glaucoma among many. The traditional enucleation technique involves the imbrication of the rectus muscles to one another with a polymethyl methacrylate (PMMA) implant. This method is cost-effective, quick, and provides reasonable motility for patients. PMMA implants are readily available and have been shown to be comparable to integrated implants in terms of efficacy and performance.^1^

The Myoconjunctival enucleation technique involves suturing the rectus muscles to the respective fornices,^2^ which adds a few extra steps and sutures. However, in many studies, this approach has been shown to provide superior motility to the prosthesis, significantly improving functional outcomes for anophthalmic patients. ^3–4^ Both aesthetic and functional motility are crucial for social acceptance and psychological well-being in such patients, providing the patient with enhanced confidence.

Despite its benefits, the Myoconjunctival technique is not widely practiced in Nepal, and there is a scarcity of published literature on its use in the region. Published studies from other countries have demonstrated that the technique provides excellent motility, low complication rates, and remains cost-effective.^3–4^ This study aims to evaluate the effectiveness of the Myoconjunctival technique within the context of our national healthcare setting, assess patient satisfaction, and promote its broader adoption across Nepal.

## Materials and methods

This non-randomised Prospective cohort Study was conducted at Tilganga Institute of Ophthalmology, Kathmandu from 30 th May 2023 to December 2023 among 35 eyes of the patients who have undergone myoconjunctival enucleation for various indications. Subjects more than 18 years undergoing unilateral enucleation with primary implant were selected, and patients with enucleation without implant, conjunctival shrinkage, adnexal deformities, and patients posted for secondary implants, patients not willing to participate and patients who could not come for scheduled follow-up were excluded from the study.

The study procedures adhered to the tenets of the Declaration of Helsinki and were conducted in compliance with the Institutional review committee of Tilganga Institute of Ophthalmology. Ethical approval was taken from Nepal health research council(NHRC) on 10^th^ may 2023 with ref no 3054. Prior to conducting the study we explained the nature of the study and the responsibility of each participant and had the freedom to withdraw from the study at any point of time. Informed written consent was taken before including in the study.Consent for usage of clinical data, photography and videography of the evaluation process was obtained from each study participant.

Based on the hospital records, in the previous year (2022), 35 cases underwent enucleation in first 6 months. Hence, 35 cases were taken as sample size and consecutive sampling was done.

Under peribulbar anaesthesia, the usual initial steps of enucleation was conducted, then each recti muscle was hooked, secured with single half of 6-0 polyglactin suture (Vicryl, Ethicon-US), released from the insertion and immediately attached to corresponding fornix by making a loop. The oblique muscle were cut, any soft tissue attachments released and finally the optic nerve was transected. Hemostasis was attained and an appropriate size poly methyl meth acrylate (PMMA) implant was inserted in the intraconal space. Implant size was calculated by subtracting 2mm from the axial length of the fellow normal eye of the patient.^5^ According to the availability, the implant size was rounded off the nearest even number. Posterior tenon and anterior sutures was sutured separately with multiple 4-0 polyglactin (Vicryl, Ethicon-US) horizontal mattress sutures. Conjunctiva was sutured using 6-0 polyglactin in continuous interlocking fashion. Appropriate sized conformer was placed and temporary tarsorrhaphy done. Any intraoperative complications and the surgery time were recorded. Enucleated specimen was sent for histopathological evaluation. All patients were given topical antibiotics and steroids in tapering frequency for six weeks of the surgery. Patients were followed up on post-operative day 1, day 7, 6 weeks and 3 months. All the surgery were done by one of the two experienced oculoplastic surgeons, HBG and MA. Prosthetic eye was given after 6 weeks of surgery. Complications on day 1, day 7, 6 weeks and 3 months were recorded. The height, width and thickness of prosthesis was measured in millimetres and volume was measured by displacement method. Hertel Ophthalmometry was done and recorded at 6 weeks.

Implant and prosthesis movement measured after 6 weeks after surgery was the primary outcome measure. A single observer measured the implant and prosthesis movement using a custom made slit-lamp device for photographic documentation, described in a study by described by Raizada et al.^6^ The device was fabricated with 2-mm scale rulers, having the dimension of 15 cm and 5 cm, respectively. The larger ruler represented the x-axis, whereas the smaller ruler represented the y-axis. The larger horizontal ruler was fixed at the center, whereas the smaller vertical ruler could be moved along the x-axis.

Under topical anesthesia, with a wire speculum in place, the center of the horizontal palpebral fissure was marked on the conjunctiva using a nontoxic color marker. The movement of the conjunctival mark in adduction, abduction, supraduction, and infraduction was photographed. The custom prosthesis then was placed in the socket. The prosthetic movement was measured in adduction, abduction, supraduction, and infraduction with photographs and real-time video documentation. The range of implant and prosthesis movement in adduction, abduction, supraduction, and infraduction from the reference point was measured. Total horizontal movement was computed by adding movements in adduction and abduction, and the total vertical movement was computed by adding movements in supraduction and infraduction.

Satisfaction score was taken from the participants as “very satisfied”, “satisfied”, “not sure”, “dissatisfied” and “very dissatisfied” ^3^ which ranged from a score of 5-1.

## Results

In this study, myoconjunctival enucleation was done in 35 eyes of 35 patients. The mean age was 31.83 ± 14.47 years, ranging from 18-70 years. Gender wise distribution of patients has been depicted in Table 1. In 14 of the 35 patients underwent surgery in the right eye and remaining 21 in the left eye.

**Table 1:** Gender wise distribution of patients.

| Gender | Frequency | Percentage |
| --- | --- | --- |
| Female | 16 | 45.7% |
| Male | 19 | 54.3% |

Figure 1 shows the number of patients with different sizes of orbital implant used. The mean implant size was 19.83 mm. The mean duration of surgery was 39.3 ± 3.76 minutes, with a range of 30 to 50 minutes.

**Figure 1:**
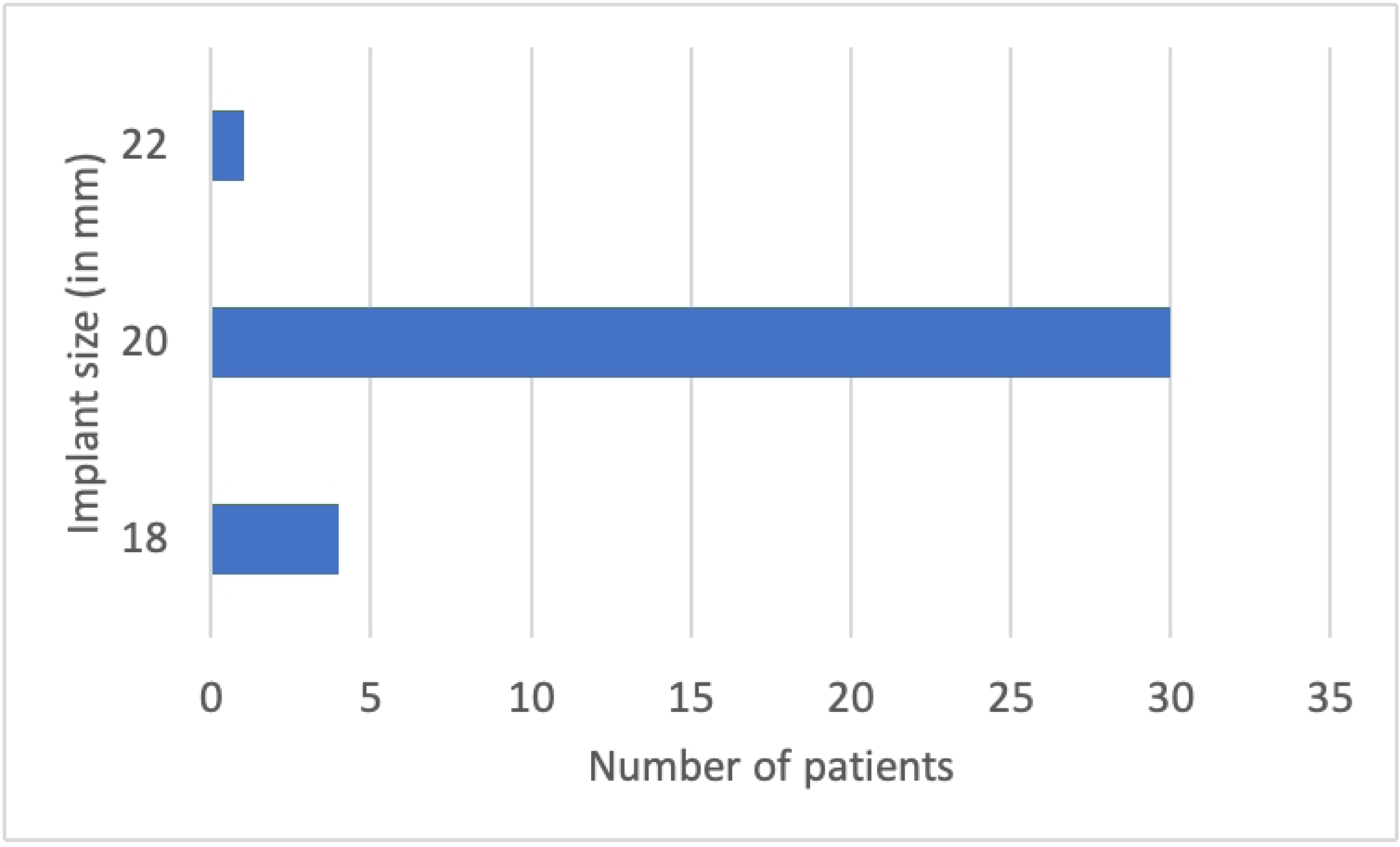
Figure depicting frequency of patient with different implant sizes.

As depicted in Table 2 and Figure 2, lateral movements (abduction & adduction) are generally better than vertical movements (elevation & depression) for both the implant and prosthesis. Prosthesis shows slightly better motility than the implant in all directions except abduction, which is equal. Depression exhibits the greatest variation (1.0–5.0 mm in prosthesis), suggesting that downward gaze is the most inconsistent across patients. The standard deviations are relatively moderate, indicating a fair consistency in motility across subjects. Overall, prosthetic motility appears superior to implant motility in most directions, particularly in depression and adduction.

**Table 2:** Implant and Prosthesis motility in different positions of gaze.

| Movement<br>(in mm) |  | Elevation | Depression | Abduction | Adduction |
| --- | --- | --- | --- | --- | --- |
| Implant | Mean | 2.343 ±<br>0.6835 | 2.257 ±<br>0.5606 | 2.743 ±<br>0.6108 | 2.714 ±<br>0.5186 |
|  | Range | 1.0- 4.0 | 1.0- 3.0 | 1.0- 4.0 | 2.0- 4.0 |
| Prosthesis | Mean | 2.457 ±<br>0.6572 | 2.629 ±<br>0.7702 | 2.743 ±<br>0.8859 | 2.829 ±<br>0.7065 |
|  | Range | 1.0- 3.0 | 1.0 -5.0 | 1.0- 4.0 | 1.0- 4.0 |

**Figure 2:**
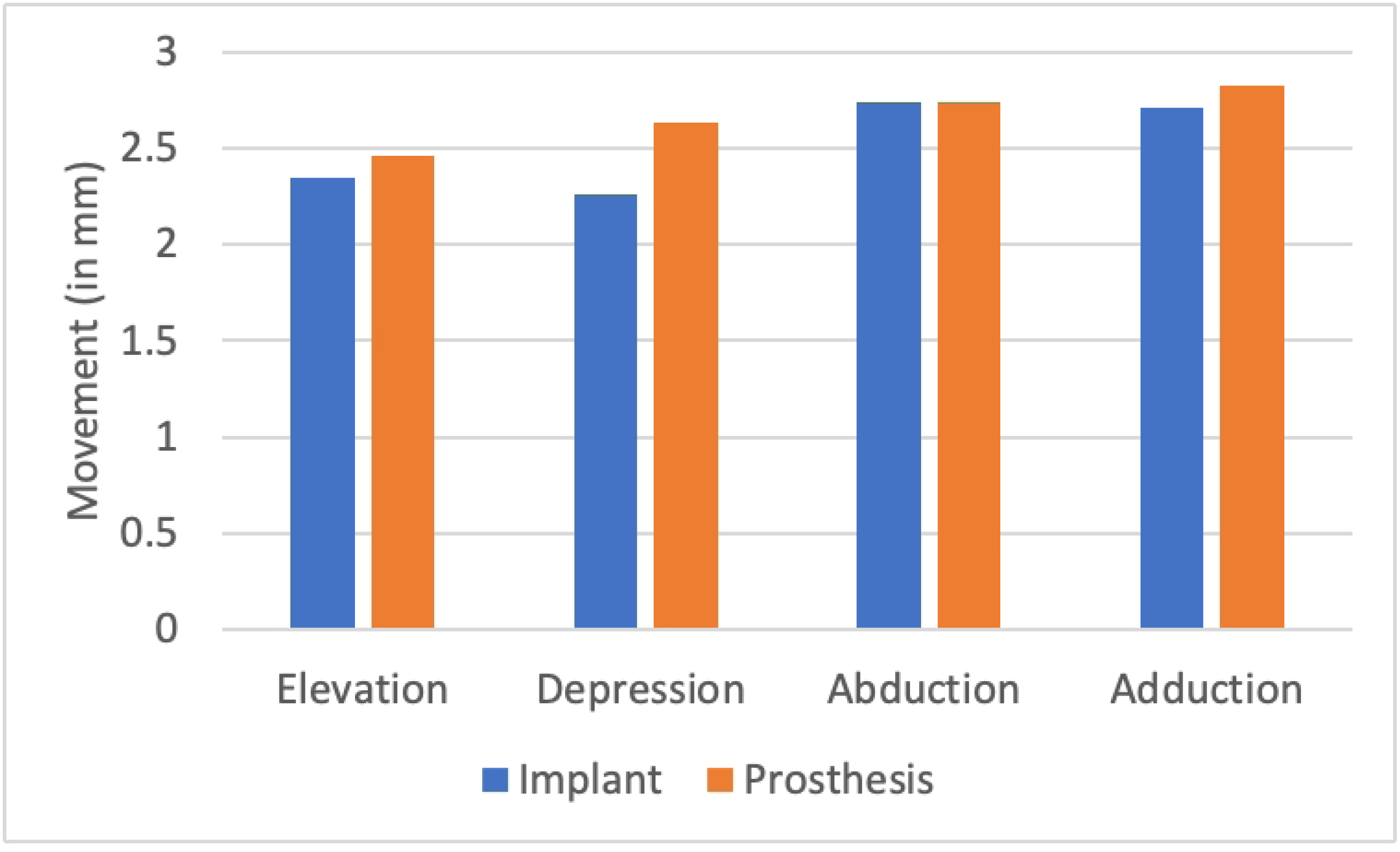
Comparative Movements of Implant and Prosthesis in Different Positions of Gaze.

Various complications observed during and after Myoconjunctival enucleation is depicted in Table 3.The most common histopathological diagnosis was phthisis bulbi (31 cases), while other conditions like endophthalmitis, perforated corneal ulcer, panophthalmitis, and staphyloma with chronic keratitis were rare, each occurring in a single case.

**Table 3:**
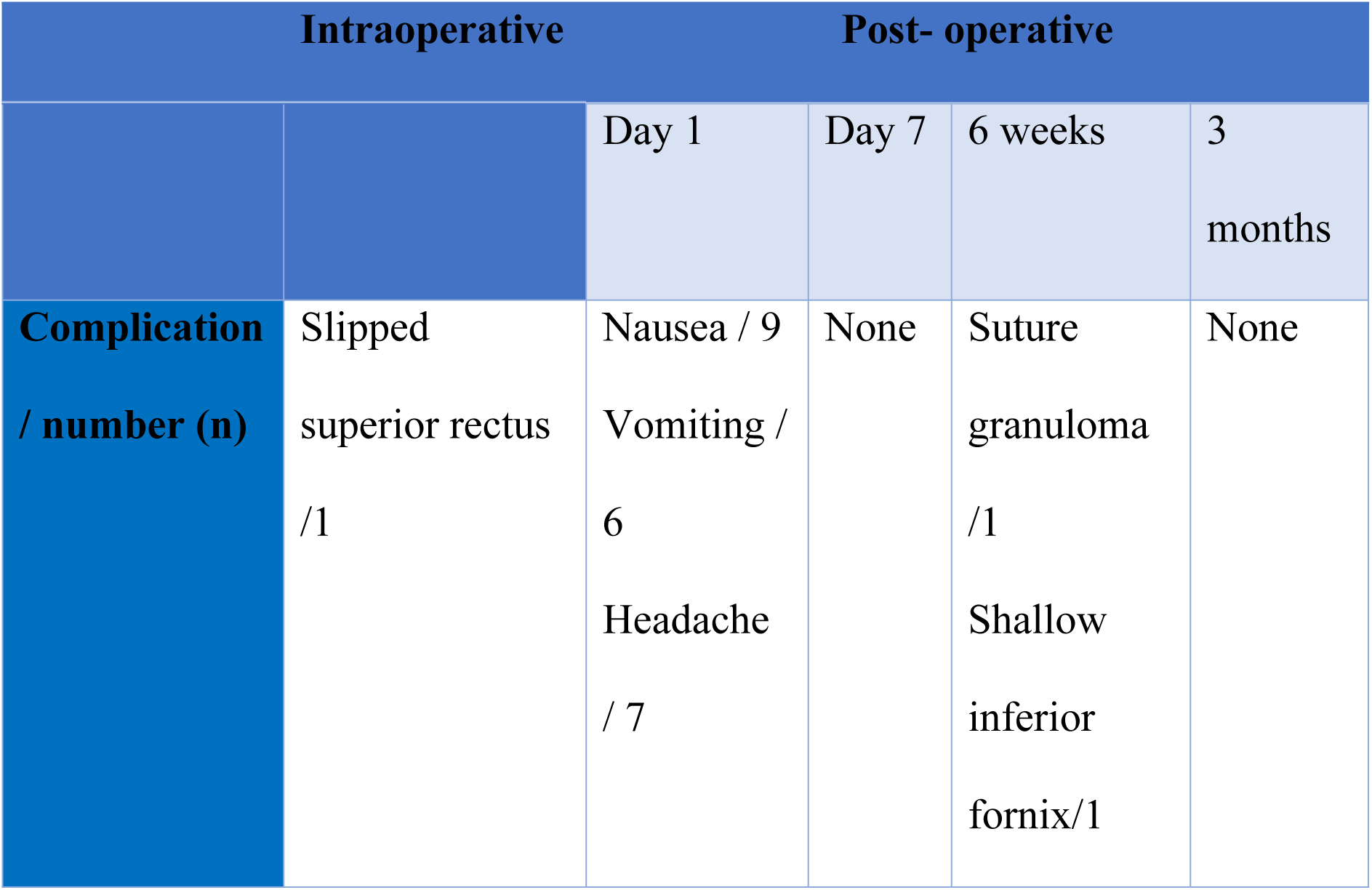
Complications observed during and after surgery at different time frames.

The majority of patients (91.94%) reported being satisfied or very satisfied with their outcomes, with a mean satisfaction score of 4.25 and a standard deviation of 0.6477 as shown in figure 3,. The scores ranged from 3 to 5, with only four patients giving a score of 3, primarily due to low motility (two patients) and a grade 1 contracted socket (one patient). This suggests that overall patient satisfaction was high, with minimal dissatisfaction.

**Figure 3:**
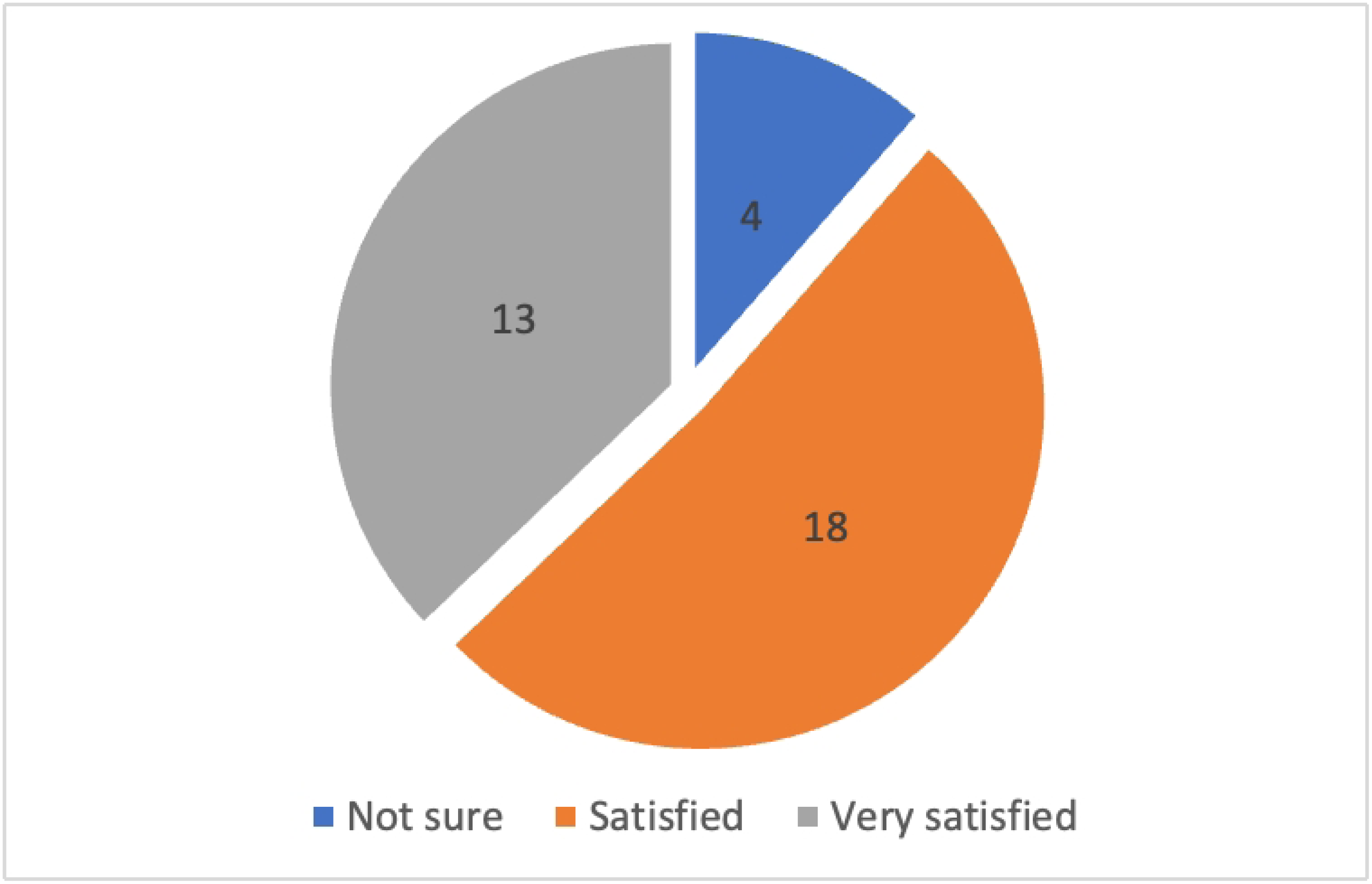
Overall patient satisfaction scores of the outcome.

## Discussion

Enucleation may be the oldest operation of ophthalmology, and as early as 2600 BC, there was a Chinese god devoted to the profession of ocularists.^7^ Enucleation is the first step to a long journey and surgeon can make a difference by the enucleation technique and the choice of implant. Replacing adequate orbital volume, maximizing prosthetic movement, comfort and aesthetic appearance should be the ultimate goal. Management of an ophthalmic socket entails type of implant, technique of surgery and ultimately the prosthesis. Orbital implant first began when Mules implanted a glass sphere after an evisceration in 1885.^7^ Various implant materials, shapes and sizes have developed since then. Orbital implant may and may not be wrapped. The rectus muscles can be imbricated (routine enucleation) to each other, attached to fornixes (Myoconjunctival technique) or attached to wrapped material.

Direct suturing of extraocular muscles to the fornixes allows movement of the muscles to be transmitted directly to the fornixes. Coston is credited with the first one to suture muscles to fornix.^8^ Since the pulling effect on the fornixes is the most important determinant of movement with the sphere implant, the motility is maximized. Nunery et al described the technique of myoconjunctival enucleation in 25 patients.^2^ The technique uses relatively cheaper spherical implant and has been shown to impart good ocular motility so is frequently used in different parts of world.^3^ In an international survey on retinoblastoma 19% surgeon preferred myoconjunctival enucleation.^9^

We used unwrapped PMMA implant in all cases. The implant and CMP/ prosthesis motility in our study was less than the study by Debraj shome et al but it is more than the routine enucleation in the same study.^3^ It is also less than the study by Asari WA et al and U Yadava et al.^10–11^ This may be due to small sample size, where a single outlier can influence the mean motility of all cases. The case with minimum motility was that of perforated corneal ulcer with conjunctival congestion.

Our average surgery time was 39.3minutes which is similar to shome et al study who had mean time of 35.2 minutes.^3^

Superior rectus was slipped in one case but retrieved and sutured. Two cases required canthotomy one case required cantholysis as added procedure. Nausea, vomiting and headache was common in first post operative day and two patients required intravenous antiemetics, all of these symptoms resolved by one week. One patient had suture granuloma at six weeks and was cauterized while one had shallow inferior fornix and fornix deepening suture was applied. There was no implant related complication in any patients. Frontalis sling surgery was done in one patient who had phthisis bulbi and ptosis from the start. Shome et al noted no complication in the myoconjunctival enucleation group,^3^ deep Superior sulcus and mild ptosis was seen in all patients with the myoconjunctival technique in Yadava et al article which was not present in our patients,^11^ conjunctivitis, implant extrusion /exposure and cellulitis were seen in 16% of myoconjunctival enucleation in an analysis of complication between myoconjunctival and conventional technique done for retinoblastoma.^4^ Custom made prosthesis was made for all patients. The length, width and thickness of all prosthesis was measured. Hertels exophthalmomemtry was done at 6 weeks. Only 4 patients had difference of 2 mm between normal eye and prosthetic eye. One had buphthalmos in both eyes so she had the highest difference of 6 mm. we used 20 mm implant size in about 85.71% which is similar to study by song et al.^12^

13 patients (37.14%) said they were very satisfied while 18 patients (51.42%) said they were satisfied while 4(11.43%) were unsure. Among the unsure patient one had less motility and one had contracted socket (shallow inferior fornix).

The patient satisfaction score arbitrarily represented the patient course of treatment and satisfaction with prosthesis.

### Limitations

It is a one-armed study. The superiority of the technique over other techniques cannot be established. Blinding/masking is not done to observer and may lead to bias. The sample size is relatively small and may not represent the general population in different setting.

### Conclusion

Myoconjunctival enucleation is a good and cheap option for imparting social motility to prosthesis with minimal complication. With minimal modification in suturing technique, the cost of suture can be brought down.

### Recommendation

Randomised control trial between routine enucleation (the most commonly performed enucleation in Nepal) and Myoconjunctival enucleation should be done. Though RCTs have been done in India, Myoconjunctival enucleation is not established among oculoplastic surgeons in the country.

### Financial support and sponsorship

Funded by cureblindness.org

## Data Availability

supporting information files

